# Uncertainty-Aware Triage of Microsatellite Instability Status in Colorectal Cancer from H&E-Stained Whole-Slide Images

**DOI:** 10.1101/2025.11.14.25340218

**Authors:** Witali Aswolinskiy, Mozzam Motiwala, Martin Paulikat, Nina Nelius, Aysel Ahadova, Patrick Adam, Dietmar Schmidt, Matthias Kloor, Christian Aichmüller

## Abstract

Microsatellite instability (MSI) is a key biomarker in colorectal cancer (CRC). Accurate distinction between MSI and microsatellite stable (MSS) tumors is critical for diagnosis, prognosis, and therapeutic decision-making. We developed an uncertainty-aware deep learning model for MSI prediction directly from H&E-stained whole-slide images. The approach is based on attention-based multiple instance learning, which can learn from slide-level labels. We extended the approach and included ensembling for more robust prediction. For training, we used 1,492 slides from three public archives and for evaluation 1,094 slides from five independent external cohorts. The approach provides predictions with confidence intervals and rejects uncertain cases. It achieved sensitivity over 97% on 59-78% of the cases, depending on the cohort. These results show that MSS cases can be identified with high confidence while uncertain cases can be rejected to avoid unreliable predictions, enabling clinically reliable MSS triage in colorectal cancer with AI.

## Introduction

Colorectal cancer (CRC) is among the most prevalent and deadly cancers worldwide, with an estimated 1.9 million new cases and over 900,000 deaths in 2022 [1]. Precision treatment of CRC requires the evaluation of genetic biomarkers, such as microsatellite instability (MSI) [2, 3], present in approximately 15% of all CRC cases. Tumors exhibiting MSI often demonstrate a favorable response to immunotherapy, while microsatellite stable (MSS) tumors are typically treated with conventional chemotherapy [4]. The determination of the MSI status is commonly achieved through laboratory techniques such as polymerase chain reaction (PCR) and immunohistochemistry (IHC). PCR is often considered the reference method and is more resource-intensive, whereas IHC is more cost-effective, but may exhibit lower sensitivity and specificity in MSI detection [5, 6].

In recent years, the application of deep learning to analyze routinely acquired hematoxylin and eosin (H&E)-stained whole-slide images (WSIs) has emerged as a promising avenue for predicting MSI status [7, 8, 9]. The main application of these tools is pre-screening or triage, aiming to reduce laboratory load. However, these methods have been less focused on reliability and decision uncertainty, which is crucial for integration into clinical practice. Erroneous MSI classification can lead to inappropriate treatment, making uncertainty-awareness a key requirement for clinical adoption. Furthermore, many proposed models lack sufficient validation across diverse populations.

In this work, we adapt Attention-Based Multiple Instance Learning (ABMIL) [10, 11] to the prediction of MSI from H&E slides in CRC. In ABMIL, the WSI is first divided into small patches, which are then encoded using a pathology vision foundation model [12]. Recent studies show that foundation models based on large vision transformers yield the highest performance in downstream tasks [13]. The ABMIL model itself is a neural network that aggregates the created patch-encodings into a slide-level representation via attention-weighted averaging and classifies them.

Methodically, our contributions are threefold. First, we show that with our chosen backbone foundation model, using the average of the register tokens rather than the classification token yields better performance for MSI classification. Second, we use only patches with the highest attention for prediction, excluding patches with low attention scores. Third, we use large ensembles of ABMIL models and include the prediction confidence interval in the final decision to increase robustness and accuracy.

We use public, multi-centric data for training and five independent external cohorts from different centers for evaluation. Overall, our approach yields high performance on previously unseen datasets while maintaining a high degree of interpretability, making it suitable for clinical practice.

## Materials and Methods

### Data

For training, we used 421 diagnostic colorectal slides from The Cancer Genome Atlas (TCGA [14, 15]), 221 slides from the Clinical Proteomic Tumor Analysis Consortium (CPTAC [16]) and 850 slides from the SurGen [17] cohort. The TCGA slides were digitized with an approximate pixel size of either 0.25 or µm, and the CPTAC slides at 0.25 µm. All training slides were converted to the generic TIFF-WSI format using either the PyVips [18] or ASAP [19] libraries. The SurGen slides were scanned with a ZEISS Axio Scan.Z1 scanner with a pixel size of 0.1112 µm and were downsampled during conversion to 0.4448 µm per pixel.

For evaluation, we used five independent external cohorts: 653 slides from the Department of Applied Tumor Biology in Heidelberg (ATB [20]), 250 slides from the Northern Ireland Biobank (NIB [21]), 99 slides from Pathology Ingolstadt (ING [22]), 47 slides from the Pathology Artificial Intelligence Platform (PAIP [23]) 2020 challenge and 45 slides from Pathology Trier (TR [24]). The ATB, ING and TR slides were scanned with a Hamamatsu NanoZoomer S210 digital slide scanner; ATB and TR with a pixel size of 0.2207 µm and ING with 0.4414 µm. The PAIP slides were scanned with an Aperio AT2 scanner with a pixel size of 0.2522 µm. The MSI status for the ATB samples was determined using PCR-based fragment length analysis [25].

A prerequisite for our approach is that slides contain sufficient, diagnostically usable tumor tissue. All evaluation slides therefore underwent a manual quality check for scanning artifacts, followed by an automated assessment of tumor sufficiency as part of our quality control procedure (see Section Method Overview). Only resection specimens were considered; biopsy slides from ING and TR were excluded in a later step due to their low sample number and quality.

A summary of all cohorts, including source, cohort size and whether they were used for training or evaluation, is provided in Table 1.

**Table 1.** Cohort composition by source, case count, MSI cases, slide count, and usage.

| Source | Cases | MSI Cases | Slides | Usage |
| --- | --- | --- | --- | --- |
| SurGen | 721 | 68 (9%) | 850 | training |
| TCGA | 416 | 61 (15%) | 421 | training |
| CPTAC | 105 | 24 (23%) | 221 | training |
| ATB | 653 | 213 (33%) | 653 | evaluation |
| NIB | 250 | 100 (40%) | 250 | evaluation |
| ING | 99 | 12 (12%) | 99 | evaluation |
| PAIP | 47 | 12 (26%) | 47 | evaluation |
| TR | 45 | 12 (27%) | 45 | evaluation |

### Method Overview

Our approach consists of three phases (see Fig. 1). The first phase is quality control based on the colorectal patch classification model from the TIA Toolbox [26, 27]. This model classifies tissue into the tissue types tumor (colorectal adenocarcinoma epithelium), tumor-associated stroma, mucus, lymphocytes, debris, muscle, fat and normal colon mucosa. We use this segmentation to determine the amount of tumor present on each slide. Preliminary experiments showed reduced model performance for slides with a small amount of tumor tissue. Therefore, we limit our approach to slides with at least 5 mm^2^ of tumor. Slides not meeting this criterion are excluded from evaluation. In the second phase, we detect tissue on the slides using both contrast-based thresholding and thresholding in the LAB color space. From the detected tissue, patches of size 224 *×* 224 px at 1 µm/px are extracted and encoded using the public foundation model H-Optimus-0 [28]. This yields a 1,536-dimensional feature vector per patch. Finally, an ensemble of ABMIL models is applied and their scores are averaged for classification into MSS, MSI, and Undetermined (i.e. inconclusive) if the prediction is uncertain. For the uncertainty estimation, we also consider the confidence interval of the ensemble prediction (see Section Ensembling and Uncertainty Estimation for more details). Each ABMIL model also outputs patch-level attention scores. We average the attention scores of these models and visualize them as heatmaps to indicate the regions most relevant for the decision.

**Figure 1.**
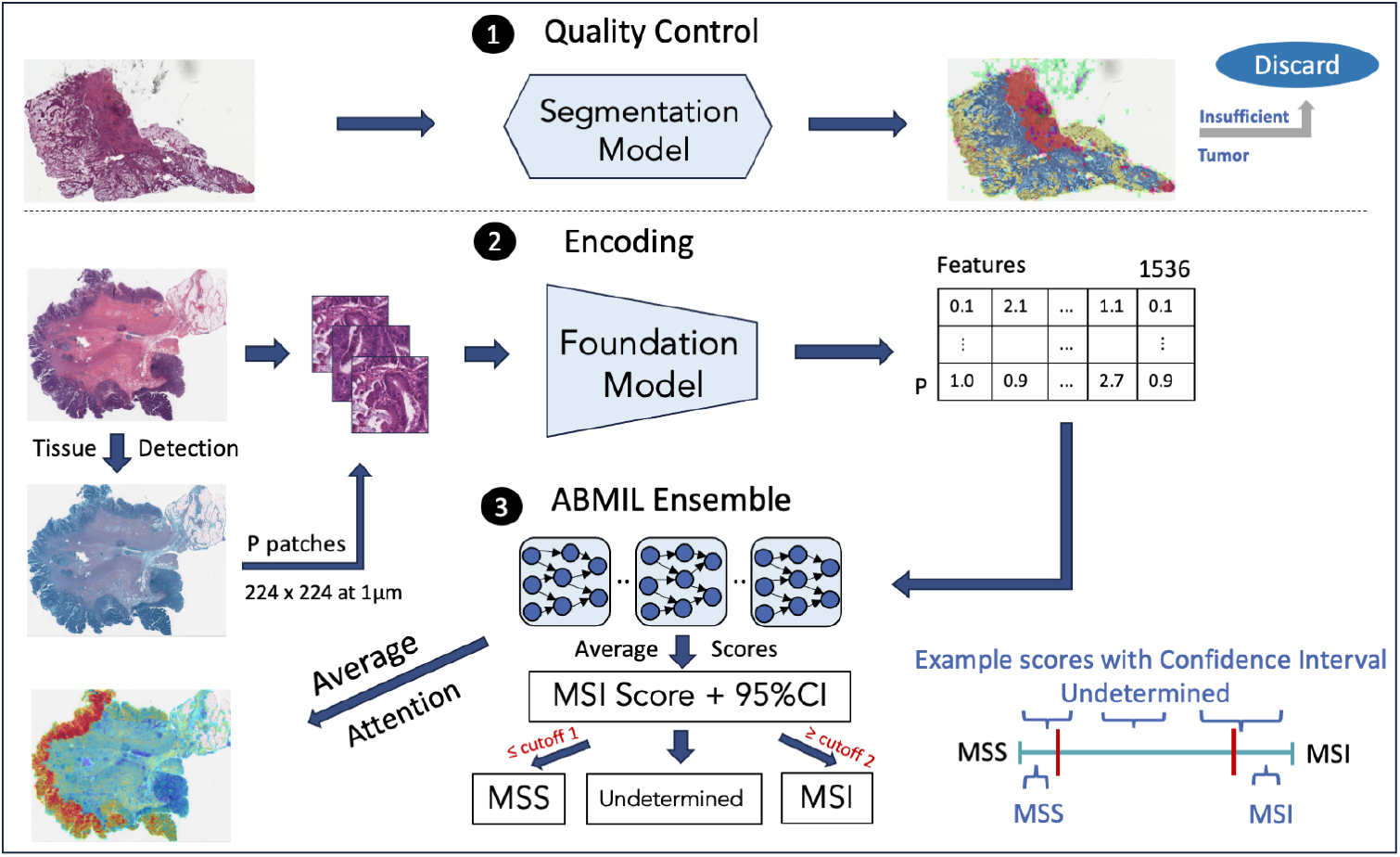
Method overview: As a preprocessing step (1), a segmentation model is applied to find and exclude slides with an insufficient amount of tumor (overlay top right; legend: blue = tumor, yellow = mucus, orange = stroma, lymphocytes = purple, green = normal/fat/muscle). Slides with sufficient tumor area are forwarded to tissue detection and encoding (2), where tissue patches of size 224 *×* 224 px are converted by the foundation model into 1,536-dimensional feature vectors. The encoded patches are processed by an ensemble of ABMIL models (3), which output an MSI score together with a 95% confidence interval based on bootstrapped ensemble predictions for each slide. Finally, slides are classified as MSS, MSI, or Undetermined according to predefined thresholds.

### Training

We split the training data into 25 independent four-fold splits and trained a model for each split resulting in an ensemble of 100 models. When multiple slides per patient were available, we ensured that a patient’s slides were included only in one fold. Each model was trained with early stopping. The training was stopped if there was no improvement in the validation area under the receiver operating characteristic curve (ROC AUC) for 15 epochs and the best (early-stopped) model was used for evaluation. As optimizer, we used AdamW [29] with learning rate 1e-4. The learning rate was reduced by a factor of 10 if there was no improvement in the validation AUC for 10 epochs.

### Layer and token choice in the foundation model

In our preliminary work [30], we showed that using intermediate layers in the foundation model can improve predictive performance compared to the final layer. Here, we additionally investigated the choice of tokens in the vision transformer foundation model. In histopathology foundation models based on vision transformers, the classification token is most often used in downstream tasks [31, 32]. Additionally, it may be concatenated with the average of the other tokens [33, 34]. Recent vision transformer architectures introduce register tokens, which may contain global image information [35]. A recent study has already shown their usefulness for out-of-distribution detection [36]. To assess which representations are most effective for MSI classification, we compared embeddings derived from different token and layer selections.

For our work, we chose the public foundation model H-Optimus-0 [28], which has demonstrated high performance on several benchmark tasks [37]. The model consists of 40 blocks with over 1.1B parameters. We evaluated the classification token and the average of the four register tokens, both in the last layer and the final LayerNorm [38] in the penultimate (39th) block.

### ABMIL and Top-K attention

The ABMIL architecture consists of three components. First, a linear projection layer reduces the dimensionality of the foundation model encodings. Next, an attention block computes an attention score for each encoded patch. These scores are normalized via the softmax function to sum to one and then used to weight patch encodings during aggregation into a slide-level representation. This representation is subsequently mapped through one or more fully connected layers to produce the final prediction. The details of our architecture are shown in the Supplementary Fig. S1.

Empirically, we observed that only a small subset of patches receives a high attention score and thus meaningfully contributes to the prediction. The remaining patches, which can number in the thousands in large WSIs, do not meaningfully contribute to the prediction and may introduce noise. To reduce this effect, we adapted the aggregation function to include only the patches with the top-*k* highest attention scores in the slide. We experimented with different values of *k* and set *k* = 10 for the final models as this choice achieved strong performance while keeping the number of evaluated patches small.

### Predicting MSS, MSI, and Undetermined

The output of the ABMIL classifier is a continuous score between 0 and 1, representing the predicted probability of MSI for a given slide. In a standard binary classification setting, the performance of the classifier is measured by the ROC-AUC, which is independent of threshold choice. For clinical triage, however, a decision threshold must be selected to distinguish MSS from non-MSS. In practice, this threshold is set low to reduce the number of false negatives (MSI cases classified as MSS). The remaining non-MSS classified cases proceed to confirmatory testing (e.g., PCR or IHC). This approach prioritizes patient safety while reducing the number of cases requiring conventional testing.

We defined this triage threshold based on the 95% true positive rate (TPR, MSI=Positive) on the validation data, so that no more than 5% of MSI cases are missed. Additionally, we evaluated a second, high-specificity threshold for confident MSI prediction, set at the 99% true negative rate (TNR, MSS=Negative), so that no more than 1% of MSS cases are incorrectly classified as MSI. This allows us to evaluate whether further reduction of the number of cases tested conventionally is possible by excluding high-confidence MSI predictions from confirmatory testing. Scores falling between these two thresholds are labeled ‘Undetermined’, requiring standard molecular testing.

### Ensembling and Uncertainty Estimation

Ensembling is a simple yet effective strategy to improve predictive performance and quantify model uncertainty. We constructed an ensemble of 100 independently trained models, whose predictions were averaged for each WSI. The effect of ensemble size on predictive performance was analyzed separately (see Supplementary Fig. S3), showing that performance plateaus at around 100 models. Additionally, we performed bootstrapping with 1,000 resamples of the model predictions to estimate the 95% confidence interval (CI). If the CI overlapped either decision threshold, the prediction was assigned to the ‘Undetermined’ category.

### External Model Comparison

For external benchmarking, we applied the Wagner et al. [39] model to all evaluation datasets. We selected this model as it represents the current state-of-the-art publicly available approach. It combines the CTransPath [40] foundation model with transformer-based feature aggregation and was trained on a multicentric cohort of over 8,000 patients. To ensure a fair model comparison, we integrated this model into our encoding pipeline without stain normalization. Ensembling was not performed, as only a single pretrained model was available.

### Online application for MSI detection

To facilitate accessibility and promote clinical translation, we implemented the proposed model as part of an online application, **SatSightDX v1.0**, which enables automatic MSI prediction from uploaded H&E-stained whole-slide images of colorectal cancer. The system is designed to be hardware- and vendor-agnostic, supporting both direct interaction via a web browser and integration with existing digital pathology infrastructures via standard interfaces such as AWS S3.

SatSightDX operates as a containerized, cloud-native application using Docker and Kubernetes for scalable and fault-tolerant deployment. After a WSI is uploaded, the platform automatically initiates the analysis pipeline, performing the quality control, tissue detection, and ensemble ABMIL inference steps described above. A structured, clinically interpretable report is generated typically within 60 minutes per slide, depending on infrastructure.

## Results

### Model comparison

We evaluated several model variants to quantify the contribution of our architectural adaptations. Our proposed model, **PB-Reg-Top10**, includes three modifications: it uses the register tokens instead of the class token, extracts them from the penultimate block rather than the last layer, and restricts classification to the top 10 attended patches.

To measure each component’s contribution (penultimate block, register tokens, top-10 selection), we additionally evaluated four ablation variants:

- *Last-CLS-All* : baseline ABMIL configuration using the class token from the last layer of H-Optimus- 0 and attention-averages all patches.
- *Last-Reg-All* : replaces the class token with register tokens, but retains the last layer and attends to all patches.
- *PB-CLS-All* : extracts the class token from the penultimate block.
- *Last-CLS-Top10* : applies the top-10 patch selection while still using the class token from the last layer.

Table 2 summarizes the evaluation results of these five model variants and the external baseline. Detailed ROC curves for **PB-Reg-Top10** are depicted in Supplementary Fig. S2.

**Table 2.** Evaluation of ensemble model variants and the public baseline across external cohorts via AUC (95% CI computed via bootstrapping with 1,000 resamples). The best result per cohort is underlined.

| Model | ATB | NIB | ING | TR | PAIP |
| --- | --- | --- | --- | --- | --- |
| Last-CLS-All | 0.934 (0.916–0.951) | 0.973 (0.955–0.988) | 0.977 (0.943–0.999) | 1.000 (1.000–1.000) | 0.983 (0.948–1.000) |
| Last-CLS-Top10 | 0.948 (0.933–0.963) | 0.974 (0.955–0.989) | 0.981 (0.950–1.000) | 1.000 (1.000–1.000) | 0.986 (0.957–1.000) |
| Last-Reg-All | 0.943 (0.926–0.959) | 0.971 (0.952–0.987) | 0.982 (0.952–1.000) | 1.000 (1.000–1.000) | 0.995 (0.979–1.000) |
| PB-CLS-All | 0.936 (0.919–0.953) | 0.974 (0.955–0.988) | 0.979 (0.949–0.999) | 1.000 (1.000–1.000) | 0.993 (0.971–1.000) |
| PB-Reg-Top10 | 0.959 (0.945–0.972) | 0.974 (0.954–0.989) | 0.983 (0.954–1.000) | 0.995 (0.977–1.000) | 0.993 (0.971–1.000) |
| Wagner et al. | 0.910 (0.885–0.934) | 0.979 (0.962–0.991) | 0.957 (0.860–0.987) | 0.941 (0.840–0.987) | 0.921 (0.814–0.991) |

Our proposed model **PB-Reg-Top10** achieved the highest AUC on ATB and ING, while performance on NIB and PAIP was comparable to the other variants. Notably, each modification yielded incremental gains on ATB, though confidence intervals overlapped across variants. The second-best variant for ATB, *Last-CLS-Top10*, performed among the best on TR and was otherwise comparable to the other variants. The external baseline by Wagner et al. achieved a slightly higher performance on NIB but markedly lower results on the other cohorts, indicating better generalization of our approach.

### Uncertainty-aware triage

Our approach predicts MSS, Undetermined and MSI, making it possible to evaluate it for triage (MSS vs non-MSS) and pre-screening (including MSI predictions). Evaluation is performed in an uncertainty-aware setting using the CI-based abstention strategy described in Ensembling and Uncertainty Estimation. The resulting metrics for triage on the external cohorts are summarized in Table 3; and the confusion matrices with all three classes are shown in Fig. 2.

**Table 3.** Evaluation results of the model across external cohorts together with 95% CI. Coverage: proportion of slides that were classified as MSS. Sensitivity: proportion of MSI slides that were not classified as MSS. NPV (Negative Predictive Value): probability that a slide predicted as MSS is truly MSS.

| Cohort | Coverage | Sensitivity | NPV |
| --- | --- | --- | --- |
| ATB | 59% (56–62) | 97.7% (95.3–99.5) | 98.6% (97.3–99.7) |
| NIB | 59% (54–65) | 98.0% (95.0–100) | 98.0% (95.0–100) |
| ING | 78% (71–85) | 100% (100–100) | 100% (100–100) |
| PAIP | 60% (47–73) | 100% (100–100) | 100% (100–100) |
| TR | 78% (64–89) | 100% (100–100) | 100% (100–100) |

**Figure 2.**
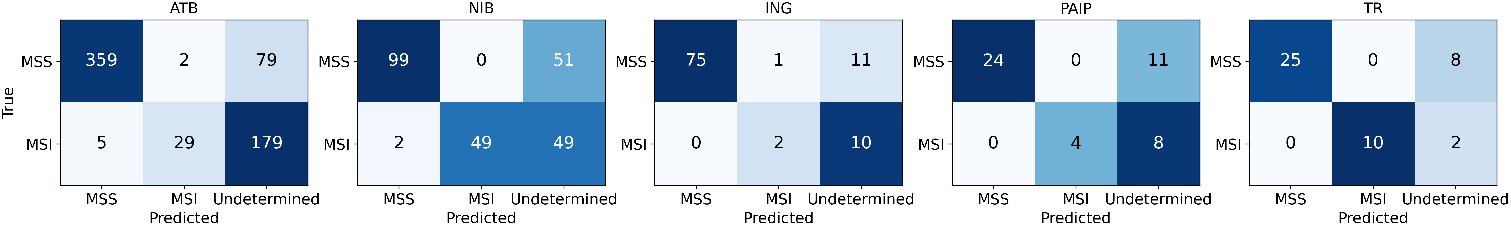
Confusion matrices of the proposed model for the external cohorts. The ground truth is shown on the y-axis (rows) and the predictions are on the x-axis (columns).

In the ATB cohort, 359 out of 440 MSS were correctly classified. Only 5 MSI cases were misclassified as MSS. In the NIB cohort, only two MSI slides were classified as MSS. Optimal results were achieved on ING, PAIP and TR with zero MSS misclassifications. Overall, high sensitivity and negative predictive value (NPV) were achieved with coverage between 59 and 78%.

To assess the impact of including prediction uncertainty into the decision process, we also evaluated the model ensemble without CI-based abstention. Coverage increased in all cohorts by (+5-10%), however, sensitivity decreased for ATB (-2.4%), with the number of false negatives increasing from 5 to 10. See Supplementary Table S1 and Fig. S4 for more details. These results indicate that utilizing prediction uncertainty may improve robustness and reliability on previously unseen cohorts.

Besides the triage scenario, we also evaluated inclusion of MSI prediction for pre-screening. However, only a relatively small fraction of MSI cases is predicted with high confidence. The coverage increases only by approx. 1%, at the cost of false negatives: 2 in the ATB cohort and 1 in the ING cohort.

### Interpretability

The ABMIL architecture provides patch-level attention scores, enabling insight into which tissue regions most strongly influenced the model’s prediction. For interpretability, we averaged the attention maps across all ensemble models. Fig. 3a shows slide thumbnails with the corresponding attention overlays, where red indicates high attention and blue low attention. Fig. 3b shows examples of the highest-attention patches from MSS and MSI slides.

**Figure 3.**
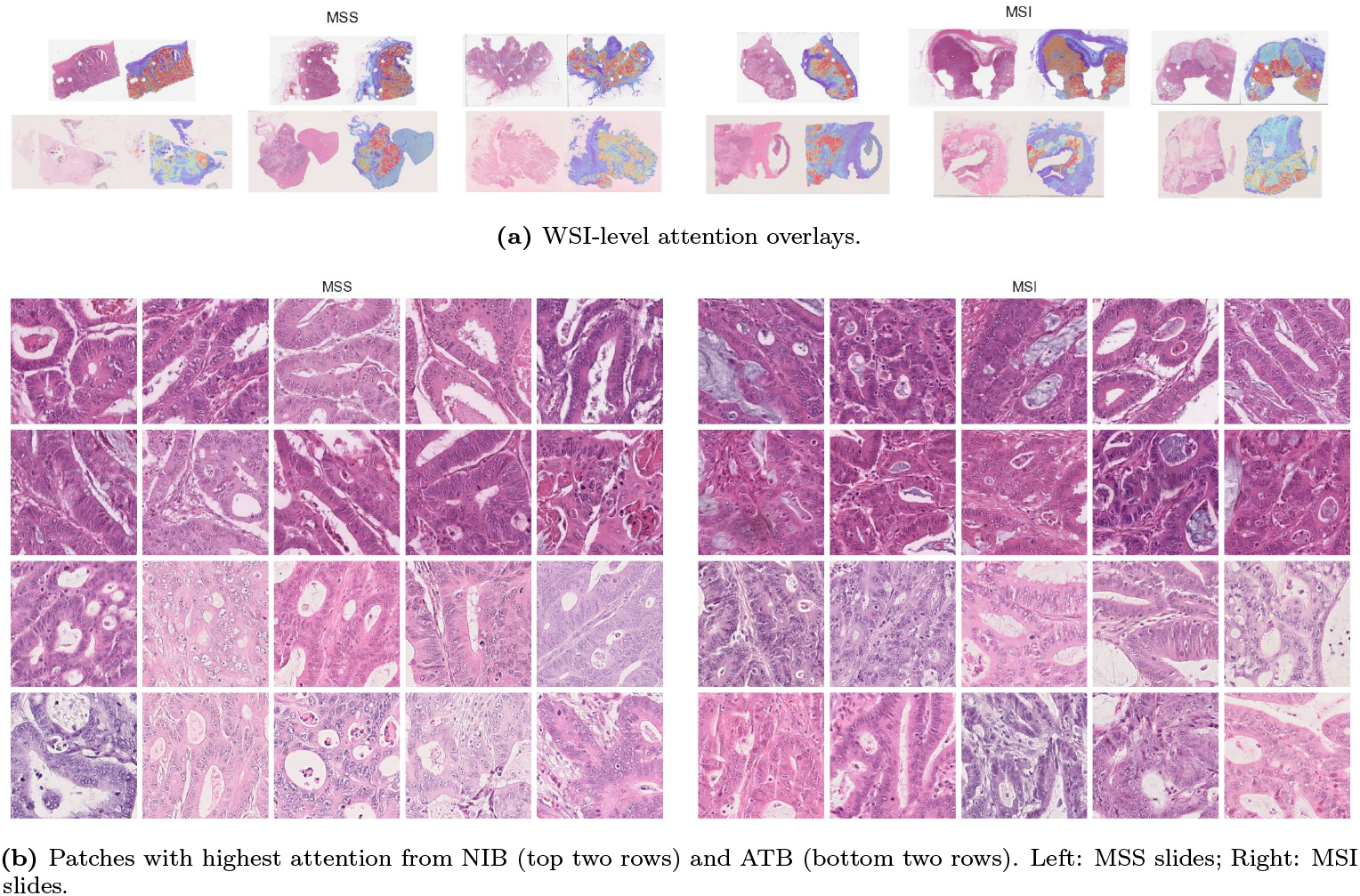
Attention-based visualizations of the proposed model. (a) Slide thumbnails with averaged attention overlays (red = high attention, blue = low attention). (b) Example patches with highest attention from MSS and MSI slides from NIB and ATB.

To quantify which tissue types contributed most to the predictions, we analyzed the tissue segmentation results of the top 1% attention patches in the ATB and NIB cohorts. On average, 59 patches (224 *×* 224 px at 1 µm/px) per slide were evaluated. The majority of attended patches corresponded to tumor regions (MSS: 85.2%, MSI: 85.9%). Other tissue types contributed less: tumor-associated stroma (MSS: 4.7%, MSI: 4.1%), necrosis/debris (MSS: 3.5%, MSI: 3.7%), mucus (MSS: 1.7%, MSI: 2.7%), muscle (MSS: 3.7%, MSI: 1.8%), normal mucosa (MSS: 0.9%, MSI: 1.2%), lymphocytes (MSS: 0.3%, MSI: 0.5%), fat (0.1% in both groups). These results indicate that the model primarily focuses on tumor areas for both classes.

Since each model in the ensemble attends to only 10 patches, we also examined the consistency of patch selection across models. For each selected patch, we measured how many models chose it. On average, only 1.7 out of 100 models selected the same patch in both the NIB and ATB cohorts. Per slide, the ensemble collectively attended to an average of 334 distinct patches. This low overlap indicates limited agreement on which patches are the most important, while the ensemble as a whole integrates information from a broader range of tissue regions, potentially enhancing robustness.

### Online application output

Each SatSightDX report includes the MSI score and corresponding confidence interval, together with visual explanations to enhance interpretability. These include tissue segmentation maps (e.g., tumor, mucus, lymphocytes) and attention heatmaps. A dedicated explanation page provides additional information for clinical users.

This deployment demonstrates that our approach can be integrated into routine digital pathology workflows, offering fast, interpretable, and uncertainty-aware MSI prediction suitable for research and potential clinical evaluation (see Fig. 4).

**Figure 4.**
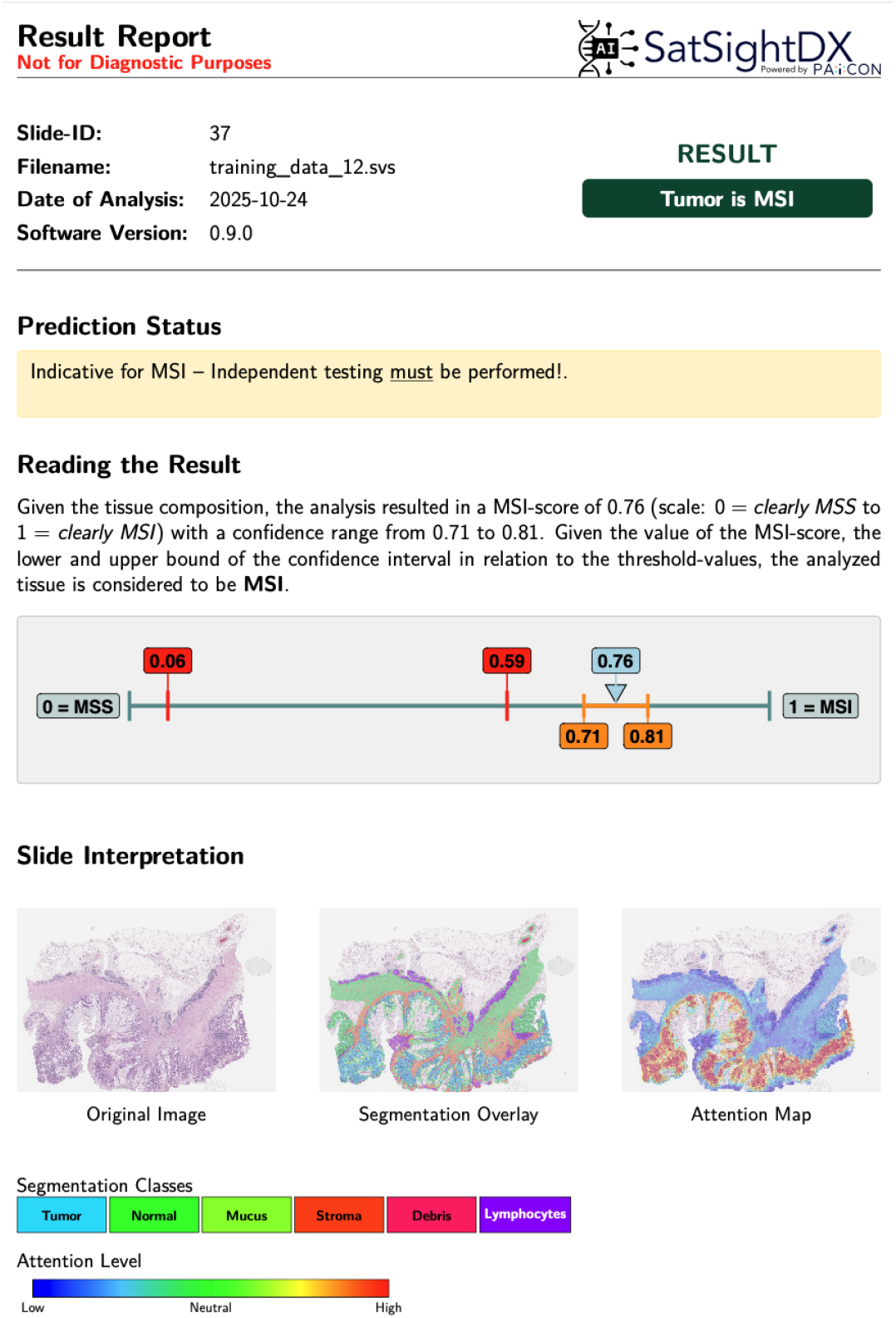
Example report from the SatSightDX online application showing MSI prediction, confidence interval, and interpretability visualizations.

## Discussion

Predicting microsatellite instability in colorectal cancer from routine H&E slides has become a common benchmark for attention-based multiple instance learning (ABMIL) models. However, many existing studies evaluate methodological variations using single models or limited repetitions, overlooking model variability. In this work, we employed large ensembles to both improve predictive performance and strengthen the validity of our ablation experiments. Although training hundreds of models is computationally demanding, inference remains fast, and the computation of confidence intervals enables an estimation of predictive uncertainty. This feature may reduce or even eliminate the need for model calibration on new cohorts. The ensemble size itself can be regarded as a hyperparameter that can be adjusted depending on the desired trade-off between computational cost and reliability.

Another key component in our approach is the number of top-attended patches. The concept of restricting prediction to the most informative instances has been explored in several MIL-based histopathology methods, including Top-1 selection or max-pooling [41], selection of both top positive and negative instances in CHOWDER [42], and differentiable top-*k* selection [43]. Our variant of top-*k* selection is intentionally simple and, similar to CHOWDER, operates within an ensemble framework. As shown in Section Interpretability, individual ensemble members attend to mostly distinct sets of patches. Consequently, the effective number of evaluated regions depends on both *k* and the ensemble size, reducing the influence of outlier regions while mitigating the inclusion of noisy or unreliable areas.

We additionally benchmarked against the publicly released model by Wagner et al. [39]. When evaluated within our preprocessing and encoding pipeline, the external baseline underperformed on four out of five cohorts compared to our variants (see Table 2). We attribute this to differences in image representations (H-Optimus-0 vs CTransPath), and more robust prediction via ensembling instead of the single released baseline model. Notably, our uncertainty-aware triage can further reduce errors by abstaining when ensemble CIs cross decision thresholds, whereas the external baseline provides point estimates only. These findings suggest that both representation choice and ensemble-based uncertainty handling may improve generalization across institutions. This comparison, however, is limited by differences in preprocessing and QC between the original setup and our pipeline.

In our evaluation, we considered two scenarios: triage, where only high-confidence MSS predictions are accepted, while the rest are forwarded to conventional testing and full pre-screening, where both high-confidence MSS and MSI predictions are used. While adding MSI predictions slightly increased overall coverage, it also introduced additional misclassifications (MSS slides incorrectly predicted as MSI). Given the clinical consequences of labeling an MSS case as MSI, even a small number of such errors is difficult to justify. Consequently, the clinically appropriate use case is safety-oriented triage that focuses on reliable identification of MSS cases while routing the remaining cases to standard molecular testing. High-confidence MSI prediction may still be valuable in research, but should not be relied upon for clinical decision-making without further prospective validation.

Our study has several limitations. First, the training and evaluation data consisted of resection specimens. Extensive validation on biopsy samples is required to confirm generalizability. Second, the quality control step relies on accurate tissue segmentation to estimate tumor area but does not assess overall specimen quality or confirm tissue type, which may present challenges. Manual quality assurance, as was done for this study, may not be feasible in clinical practice. Third, evaluation was conducted at the slide level; patient-level aggregation strategies were not explored and may affect clinical workflows when multiple slides per patient are available. Finally, while our approach does not require cohort-specific calibration, performance could potentially be improved through calibration if sufficient data is available.

The proposed approach was applied here to uncertainty-aware MSS triage for colorectal cancer, but can be readily extended to other cancer types and to the prediction of additional molecular or genetic targets. Future work will also incorporate automated quality assessment and artifact detection to further enhance clinical robustness and reduce the need for manual quality assurance, representing an important step toward broadly applicable, uncertainty-aware precision diagnostics in computational pathology.

## Data Availability

The TCGA slides are available at https://portal.gdc.cancer.gov. The CPTAC slides are available at https://www.cancerimagingarchive.net/collection/cptac-coad. Their MSI status was retrieved from cBioPortal: https://www.cbioportal.org/study/summary?id=coad_cptac_2019. The SurGen cohort is available at https://www.ebi.ac.uk/biostudies/bioimages/studies/S-BIAD1285.

The remaining data used to evaluate the models cannot be shared due to data agreement limitations.

## Code Availability

H-Optimus-0 is available from https://huggingface.co/bioptimus/H-optimus-0. Our model can be tried out after registration via a web application: https://satsight-dx.paicon.com

## Acknowledgements

We would like to acknowledge Jonathan Doerre, Ricarda Mehr, Gizem Erisen and Hugo Montémont for excellent technical assistance and scientific discussions.

## Author contributions statement

W.A. conducted the experiments, analyzed the results and wrote the manuscript. M.M., M.P., N.N., A.A., P.A., D.S. and M.K. assisted with data acquisition and preprocessing. C.A. supervised the experiments. All authors reviewed the manuscript.

## Ethics approval and consent to participate

This study is a retrospective analysis of pseudonymized whole-slide images from multiple colorectal cancer cohorts. All data was collected in accordance with local ethical and legal requirements and anonymized prior to analysis. The utilization of ATB samples was approved by the institutional Ethics Committee of Heidelberg University (S-348/2023); other sites obtained equivalent local approvals. External data was processed on-site at the corresponding institutions.

## Declaration of generative AI and AI-assisted technologies in the manuscript preparation process

During the preparation of this work, the authors used ChatGPT 5.1 to assist with minor phrasing improvements and grammatical refinement. All content generated with this tool was reviewed, revised, and approved by the authors, who take full responsibility for the final manuscript.

## References

[1] F. Bray, M. Laversanne, H. Sung, J. Ferlay, R. L. Siegel, I. Soerjomataram, A. Jemal, Global cancer statistics 2022: Globocan estimates of incidence and mortality worldwide for 36 cancers in 185 countries, CA: a cancer journal for clinicians 74 (3) (2024) 229–263.

[2] E. Vilar, S. B. Gruber, Microsatellite instability in colorectal cancer—the stable evidence, Nature reviews Clinical oncology 7 (3) (2010) 153–162.

[3] C. R. Boland, A. Goel, Microsatellite instability in colorectal cancer, Gastroenterology 138 (6) (2010) 2073–2087.

[4] M. J. Overman, R. McDermott, J. L. Leach, S. Lonardi, H.-J. Lenz, M. A. Morse, J. Desai, A. Hill, M. Axelson, R. A. Moss, et al., Nivolumab in patients with metastatic dna mismatch repair-deficient or microsatellite instability-high colorectal cancer (checkmate 142): an open-label, multicentre, phase 2 study, The lancet oncology 18 (9) (2017) 1182–1191.

[5] C. R. Boland, A. Goel, Microsatellite instability in colorectal cancer, Gastroenterology 138 (6) (2010) 2073–2087.

[6] J. Shia, Immunohistochemistry versus microsatellite instability testing for screening colorectal cancer patients at risk for hereditary nonpolyposis colorectal cancer syndrome: part i. the utility of immunohistochemistry, The Journal of molecular diagnostics 10 (4) (2008) 293–300.

[7] J. N. Kather, A. T. Pearson, N. Halama, D. Jäger, J. Krause, S. H. Loosen, A. Marx, P. Boor, F. Tacke, U. P. Neumann, et al., Deep learning can predict microsatellite instability directly from histology in gastrointestinal cancer, Nature medicine 25 (7) (2019) 1054–1056.

[8] M. Bilal, S. E. A. Raza, A. Azam, S. Graham, M. Ilyas, I. A. Cree, D. Snead, F. Minhas, N. M. Rajpoot, Development and validation of a weakly supervised deep learning framework to predict the status of molecular pathways and key mutations in colorectal cancer from routine histology images: a retrospective study, The Lancet Digital Health 3 (12) (2021) e763–e772.

[9] A. Echle, N. T. Rindtorff, T. J. Brinker, T. Luedde, A. T. Pearson, J. N. Kather, Deep learning in cancer pathology: a new generation of clinical biomarkers, British journal of cancer 124 (4) (2021) 686–696.

[10] M. Ilse, J. Tomczak, M. Welling, Attention-based deep multiple instance learning, in: International conference on machine learning, PMLR, 2018, pp. 2127–2136.

[11] M. Y. Lu, D. F. Williamson, T. Y. Chen, R. J. Chen, M. Barbieri, F. Mahmood, Data-efficient and weakly supervised computational pathology on whole-slide images, Nature biomedical engineering 5 (6) (2021) 555–570.

[12] Y. Zhang, Z. Gao, K. He, C. Li, R. Mao, From patches to wsis: A systematic review of deep multiple instance learning in computational pathology, Information Fusion (2025) 103027.

[13] G. Campanella, S. Chen, M. Singh, R. Verma, S. Muehlstedt, J. Zeng, A. Stock, M. Croken, B. Veremis, A. Elmas, et al., A clinical benchmark of public self-supervised pathology foundation models, Nature Communications 16 (1) (2025) 3640.

[14] Cancer Genome Atlas Network and others, Comprehensive molecular characterization of human colon and rectal cancer, Nature 487 (7407) (2012) 330.

[15] Y. Liu, N. S. Sethi, T. Hinoue, B. G. Schneider, A. D. Cherniack, F. Sanchez-Vega, J. A. Seoane, F. Farshidfar, R. Bowlby, M. Islam, et al., Comparative molecular analysis of gastrointestinal adenocarcinomas, Cancer cell 33 (4) (2018) 721–735.

[16] N. J. Edwards, M. Oberti, R. R. Thangudu, S. Cai, P. B. McGarvey, S. Jacob, S. Madhavan, K. A. Ketchum, The cptac data portal: a resource for cancer proteomics research, Journal of proteome research 14 (6) (2015) 2707–2713.

[17] C. Myles, I. H. Um, C. Marshall, D. Harris-Birtill, D. J. Harrison, Surgen: 1020 h&e-stained whole-slide images with survival and genetic markers, GigaScience 14 (2025) giaf086.

[18] J. Cupitt, Pyvips (version 2.2.1), https://libvips.github.io/pyvips/, released June 12, 2022 (2022).

[19] G. Litjens, Asap (version 2.2, nightly), https://github.com/computationalpathologygroup/ASAP/releases/tag/ASAP-2.2-(Nightly) (mAug. 2023).

[20] Department of Applied Tumor Biology, Institute of Pathology, University Hospital Heidelberg, Dataset from the department of applied tumor biology, university hospital heidelberg, accessed 2025 (2025).

[21] C. Lewis, S. McQuaid, P. Clark, T. McGuigan, P. Murray, C. Greene, B. Coulter, K. Mills, J. James, The northern ireland biobank: a cancer focused repository of science, Open Journal of Bioresources 5 (2018) 9.

[22] Pathologie Ingolstadt, Dataset from pathologie ingolstadt, accessed 2025 (2025).

[23] K. Kim, K. Lee, S. Cho, D. U. Kang, S. Park, Y. Kang, H. Kim, G. Choe, K. C. Moon, K. S. Lee, et al., Paip 2020: Microsatellite instability prediction in colorectal cancer, Medical Image Analysis 89 (2023) 102886.

[24] Pathologie Trier, Dataset from pathologie trier, accessed 2025 (2025).

[25] P. Findeisen, M. Kloor, S. Merx, C. Sutter, S. M. Woerner, N. Dostmann, A. Benner, B. Dondog, M. Pawlita, W. Dippold, et al., T25 repeat in the 3’ untranslated region of the casp2 gene: a sensitive and specific marker for microsatellite instability in colorectal cancer, Cancer research 65 (18) (2005) 8072–8078.

[26] J. Pocock, S. Graham, Q. D. Vu, M. Jahanifar, S. Deshpande, G. Hadjigeorghiou, A. Shephard, R. M. S. Bashir, M. Bilal, W. Lu, D. Epstein, F. Minhas, N. M. Rajpoot, S. E. A. Raza, TIAToolbox as an end-to-end library for advanced tissue image analytics, Communications Medicine 2 (1) (2022) 120. doi:10.1038/s43856-022-00186-5. URL https://www.nature.com/articles/s43856-022-00186-5

[27] J. N. Kather, N. Halama, A. Marx, 100,000 histological images of human colorectal cancer and healthy tissue, (No Title) (2018).

[28] C. Saillard, R. Jenatton, F. Llinares-López, Z. Mariet, D. Cahané, E. Durand, J.-P. Vert, H-optimus-0 (2024). URL https://github.com/bioptimus/releases/tree/main/models/h-optimus/v0

[29] Loshchilov, F. Hutter, Decoupled weight decay regularization, arXiv preprint 1711.05101 (2017).

[30] W. Aswolinskiy, M. Paulikat, C. Aichmueller, Impact of layer selection in histopathology foundation models on downstream task performance, in: Medical Imaging with Deep Learning, 2024.

[31] R. J. Chen, T. Ding, M. Y. Lu, D. F. Williamson, G. Jaume, A. H. Song, B. Chen, A. Zhang, D. Shao, M. Shaban, et al., Towards a general-purpose foundation model for computational pathology, Nature medicine 30 (3) (2024) 850–862.

[32] H. Xu, N. Usuyama, J. Bagga, S. Zhang, R. Rao, T. Naumann, C. Wong, Z. Gero, J. González, Y. Gu, et al., A whole-slide foundation model for digital pathology from real-world data, Nature 630 (8015) (2024) 181–188.

[33] E. Vorontsov, A. Bozkurt, A. Casson, G. Shaikovski, M. Zelechowski, K. Severson, E. Zimmermann, Hall, N. Tenenholtz, N. Fusi, et al., A foundation model for clinical-grade computational pathology and rare cancers detection, Nature medicine 30 (10) (2024) 2924–2935.

[34] A. Filiot, N. Dop, O. Tchita, A. Riou, R. Dubois, T. Peeters, D. Valter, M. Scalbert, C. Saillard, G. Robin, et al., Distilling foundation models for robust and efficient models in digital pathology, arXiv preprint 2501.16239 (2025).

[35] T. Darcet, M. Oquab, J. Mairal, P. Bojanowski, Vision transformers need registers, arXiv preprint 2309.16588 (2023).

[36] S. Yellapragada, K. Thopalli, V. Narayanaswamy, W. Sakla, Y. Liu, Y. Mubarka, D. Samaras, J. J. Thiagarajan, Leveraging registers in vision transformers for robust adaptation, in: ICASSP 2025-2025 IEEE International Conference on Acoustics, Speech and Signal Processing (ICASSP), IEEE, 2025, pp. 1–5.

[37] G. Jaume, P. Doucet, A. Song, M. Y. Lu, C. Almagro Pérez, S. Wagner, A. Vaidya, R. Chen, D. Williamson, A. Kim, et al., Hest-1k: A dataset for spatial transcriptomics and histology image analysis, Advances in Neural Information Processing Systems 37 (2024) 53798–53833.

[38] L. Ba, J. R. Kiros, G. E. Hinton, Layer normalization, arXiv preprint 1607.06450 (2016).

[39] S. J. Wagner, D. Reisenbüchler, N. P. West, J. M. Niehues, J. Zhu, S. Foersch, G. P. Veldhuizen, P. Quirke, H. I. Grabsch, P. A. van den Brandt, et al., Transformer-based biomarker prediction from colorectal cancer histology: A large-scale multicentric study, Cancer cell 41 (9) (2023) 1650–1661.

[40] X. Wang, S. Yang, J. Zhang, M. Wang, J. Zhang, W. Yang, J. Huang, X. Han, Transformer-based unsupervised contrastive learning for histopathological image classification, Medical image analysis 81 (2022) 102559.

[41] G. Campanella, M. G. Hanna, L. Geneslaw, A. Miraflor, V. Werneck Krauss Silva, K. J. Busam, E. Brogi, V. E. Reuter, D. S. Klimstra, T. J. Fuchs, Clinical-grade computational pathology using weakly supervised deep learning on whole slide images, Nature medicine 25 (8) (2019) 1301–1309.

[42] P. Courtiol, E. W. Tramel, M. Sanselme, G. Wainrib, Classification and disease localization in histopathology using only global labels: A weakly-supervised approach, arXiv preprint 1802.02212 (2018).

[43] S. Kapse, P. Pati, S. Das, J. Zhang, C. Chen, M. Vakalopoulou, J. Saltz, D. Samaras, R. R. Gupta, P. Prasanna, Si-mil: Taming deep mil for self-interpretability in gigapixel histopathology, in: Proceedings of the IEEE/CVF Conference on Computer Vision and Pattern Recognition, 2024, pp. 11226–11237.

