## Supplementary Material for "Uncertainty-Aware Triage of Microsatellite Instability Status in Colorectal Cancer from H&E-Stained Whole-Slide Images"

### Additional Methods

The proposed ABMIL architecture consists of three main components, implemented with fully connected layers, as illustrated in Fig. S1. The first projection layer maps the 1,536-dimensional patch embeddings to 512 dimensions and applies dropout with a rate of 25%. The subsequent attention block contains two parallel branches with `tanh` and `sigmoid` activations, respectively. Each branch reduces the input to 256 dimensions, after which the outputs are combined, and a `softmax` is applied to the top-10 attention scores. All remaining attention values are set to zero.

The resulting attention vector is multiplied with the corresponding 512-dimensional patch embeddings (from the projection layer) to form a single 512-dimensional slide-level representation. This representation is then mapped through the final classification layer to two output dimensions (MSS and MSI), followed by a `softmax` activation to obtain probability scores. Overall, this ABMIL classifier contains approximately one million trainable parameters.

The main difference from the standard ABMIL architecture lies in the top-10 attention selection, which restricts the aggregation to the most informative patches.

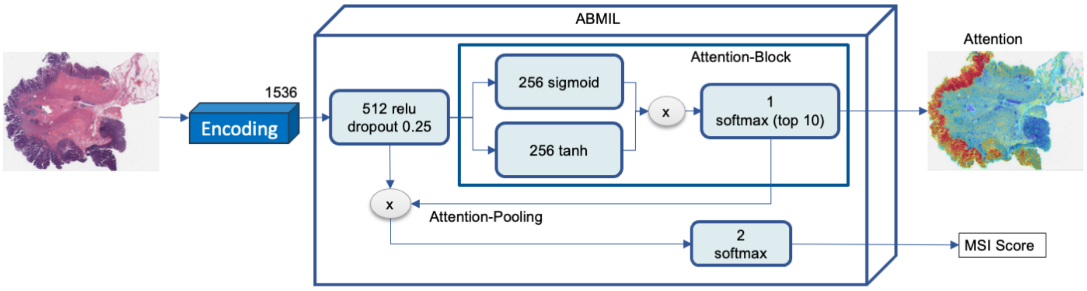


**Figure S1:** Visualization of the proposed ABMIL model architecture. The model includes a dimensionality reduction layer, an attention mechanism operating on the top-10 attended patches, and a final classification layer for MSI prediction.

### Additional Results

#### *ROC-Plots*

Fig. S2 shows the ROC-curves of the **PB-Reg-Top10** model per evaluation cohort.

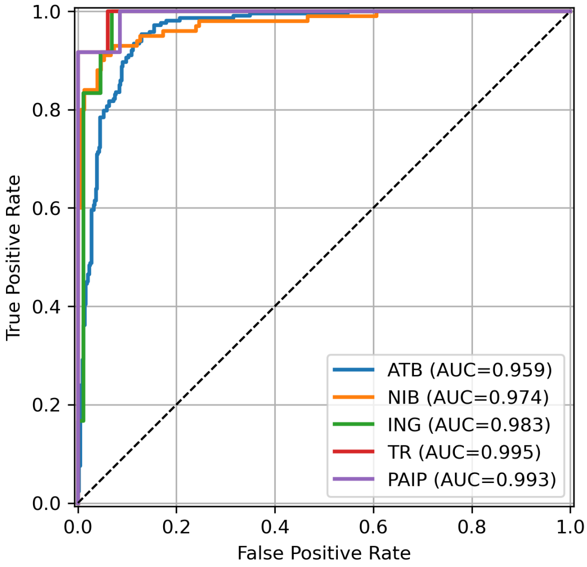


**Figure S2:** ROC-curves with AUC values for the evaluation cohorts.

*Effect of ensemble size*

We empirically determined the number of models to use in the ensemble by analyzing the impact of the number of models on predictive performance. To this end, we trained 200 independent models and evaluated ensemble performance using bootstrapping with 1,000 resamples. For each ensemble size, models were sampled with replacement, and the mean AUC was computed across resamples.

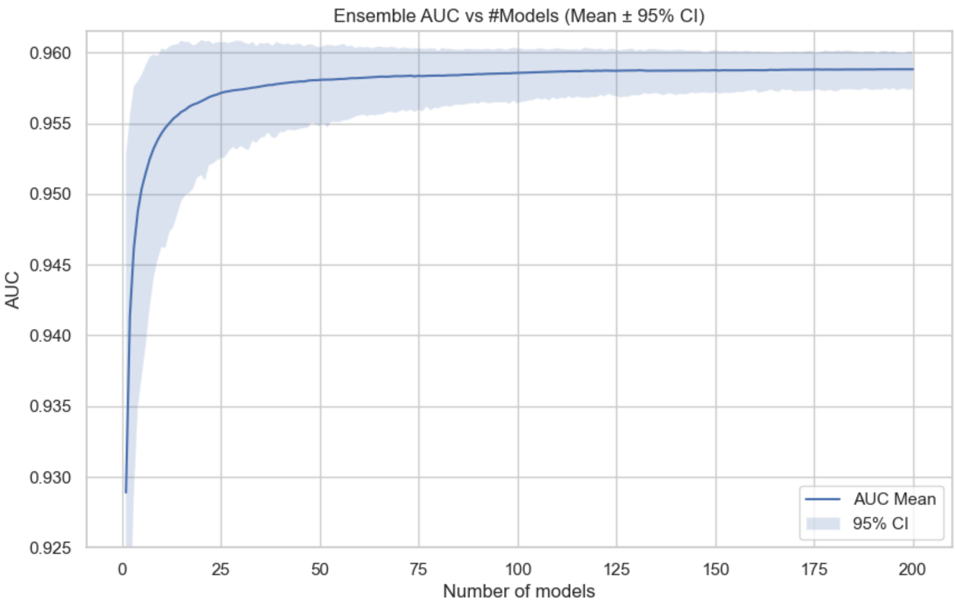


**Figure S3:** Impact of ensemble size on MSI prediction performance. The mean AUC was computed by bootstrapping with replacement across 1,000 resamples for each ensemble size.

As shown in Fig. S3, performance increases with the number of models in the ensemble but begins to plateau at approximately 100 models. Based on this observation, all experiments reported in the main text use ensembles of 100 models, which balance high performance and computational efficiency.

*Evaluation without confidence interval–based abstention*

To assess the impact of uncertainty-aware abstention, we evaluated the model ensemble without excluding predictions whose 95% confidence interval (CI) crossed the decision thresholds. Table S1 summarizes the corresponding performance metrics, and Fig. S4 shows the resulting confusion matrices.

| Cohort | Coverage | Sensitivity | NPV |
|---|---|---|---|
| ATB | 65% (62–67) | 95.3% (92.5–97.7) | 97.4% (95.9–98.8) |
| NIB | 64% (59–70) | 98.0% (95.0–100) | 98.2% (95.4–100) |
| ING | 83% (76–89) | 100% (100–100) | 100% (100–100) |
| PAIP | 70% (59–83) | 100% (100–100) | 100% (100–100) |
| TR | 84% (73–93) | 100% (100–100) | 100% (100–100) |

**Table S1:** Evaluation results of the model across external cohorts together with 95% CI. Coverage: proportion of slides that were classified as MSS. Sensitivity: proportion of MSI slides that were not classified as MSS. NPV (Negative Predictive Value): probability that a slide predicted as MSS is truly MSS.

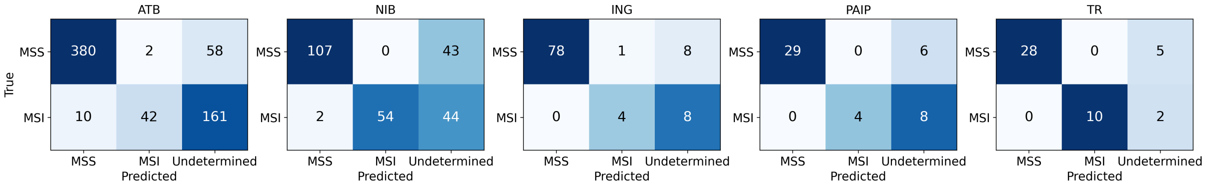


**Figure S4:** Confusion matrices of the model ensemble evaluated without CI-based abstention. The ground truth is shown on the y-axis (rows) and predictions on the x-axis (columns).

Compared to the uncertainty-aware evaluation, omitting CI-based abstention increased coverage across all cohorts. However, it reduced sensitivity in the ATB cohort. These results indicate that including uncertainty into the decision process may improve robustness on previously unseen data at the cost of lower coverage.
